# Seroprevalence and epidemiological characteristics of immunoglobulin M and G antibodies against SARS-CoV-2 in asymptomatic people in Wuhan, China: a cross-sectional study

**DOI:** 10.1101/2020.06.16.20132423

**Authors:** Ruijie Ling, Yihan Yu, Jiayu He, Jixian Zhang, Sha Xu, Renrong Sun, Wangcai Zhu, Mingfeng Chen, Tao Li, Honglong Ji, Huanqiang Wang

**Affiliations:** Integrated Chinese & Western Medicine, Hubei Provincial Hospital, Wuhan, China; National Institute of Occupational Health and Poison Control, Chinese Center for Disease Control and Prevention, Beijing, China; Department of Cellular and Molecular Biology, University of Texas Health Science Centre, Tyler, USA

**Keywords:** COVID-19, SARS-CoV-2, serological testing, IgG, IgM, seroprevalence, computed tomography

## Abstract

**Objectives:** Population screening for IgG and IgM against SARS-CoV-2 was initiated on March 25 and was open to all residents of Wuhan who were symptom-free. All ages with no fever, headache or other symptoms of COVID-19 among residents in Wuhan were included.

**Methods:** This study adopted a cross-sectional study. Pearson Chi-square test, T-test, and Mann-Whitney test were used in comparison between different groups. To correct the effects of gender and age, the seroprevalence of IgM positivity, IgG positivity, and IgM and/or IgG positivity were standardized according to the gender and age-specific population of Wuhan in 2017.

**Results:** The seroprevalence of IgG and IgM standardized for age and gender in Wuhan showed a downward trend. No significant correlation was observed between the seroprevalence of IgG and the different age groups. The seroprevalence was significantly higher for females than males (x^2^ =35.702, p < 0.001), with an odds ratio of 1.36 (95% CI: 1.24–1.48). A significant difference was seen in the seroprevalence of IgG among people from different geographic areas and different types of workplaces (respectively, x^2^ = 42.871, p < 0.001 and x^2^ = 202.43, p < 0.001). The IgG antibody-positive cases had a greater number of abnormalities in CT imaging than IgG-negative cases (30.7% vs 19.7%).

**Conclusions:** Our work found the reported number of confirmed patients in Wuhan only represents a small proportion of the total number of infections. There was a significant aggregation of asymptomatic infections in individuals from some occupations, and based on CT and laboratory findings, some damage may have occurred in asymptomatic individuals positive for IgG antibody.

Strengths and limitations of this study
- This study has the important feature of having been designed as repeated five-day serosurveys, which allowed for the monitoring of seroprevalence progression.
- This study applied scientific statistical methods accounting for the demographic structure of the general population and imperfect diagnostic tests to estimate seroprevalence in the overall population.
- This study had selection bias since the analyzed medical records were based on examinees directed by their work units.
- People under the age of 19 and over age 65 were too few to be fully covered in analyses.

## INTRODUCTION

Coronavirus disease 2019 (COVID-19) caused by Severe Acute Respiratory Syndrome Coronavirus 2 (SARS-CoV-2) is a pandemic that has spread around the globe in the early months of 2020. [1-2] Quantitative reverse transcription-polymerase chain reaction (RT-PCR) analysis for SARS-CoV-2 is the gold standard for COVID-19 diagnosis.[3-4] However, epidemiological surveillance of confirmed COVID-19 cases captures only a portion of all infections because the clinical spectrum of SARS-CoV-2 infection ranges from mild to the onset of severe acute respiratory distress syndrome.[5] Many patients can be asymptomatic, greatly increasing the uncertainty of diagnosis.[6] We know that SARS-CoV-2-specific antibodies appear around 3–10 days after infection, with IgM appearing first and IgG following around 14 days after infection.[7] Even after the symptoms of infection disappear, IgG can still show seropositivity.[8] Therefore, SARS-CoV-2 serological assays are a good supplementary method to verify nucleic acid detection.[9]

A series of serological surveys of SARS-CoV-2 have been reported.[10-17] However, many of them are small or based on non-random sampling of participants, some of them focusing on health-care workers or blood donors, and thus cannot provide precise estimates of seroprevalence by age group in the general population.[18] Additionally, most of these studies do not take into account the sensitivity or specificity of the antibody tests, or do not adjust for the age and gender of the general population, which decreases the accuracy of the estimation.

The COVID-19 outbreak was first reported in the city of Wuhan, Hubei Province, China.[19] As Wuhan accepted the timely prevention strategies of lockdown for curbing population flow, it effectively controlled the spread of the pandemic. After 76 days of lockdown in Wuhan, the city reopened again on April 8, 2020. The residents have been allowed to resume working since that day if they met a set of COVID-19-associated tests, including SARS-CoV-2 nucleic acid tests (NAT) of nasopharyngeal swabs, chest CT scans or a SARS-CoV-2-specific serological test.[20-21] Serological surveys are the best tool to determine the spread of an infectious disease, particularly in the presence of asymptomatic cases.[22] These tests provide a unique opportunity to study the number of SARS-CoV-2 infections in the asymptomatic general population.

We choose a general hospital in Jianghan District near the Huanan Seafood Wholesale Market to conduct a serological survey of those people resuming work and other inhabitants in Wuhan using a validated assay for IgM and IgG antibodies against SARS-CoV-2. We chose this area because the Jianghan District was the early epicenter of the COVID-19 outbreak in China and many of the earliest COVID-19 cases were linked to the Huanan Seafood Wholesale Market in the same district of Wuhan.[23] We selected a serology test that has been fully validated using serum from confirmed COVID-19-infected individuals and well-characterized reference samples. In this study, we analyzed 18,712 medical examination records at this hospital including IgM and IgG tests for SARS-CoV-2 antibodies and other tests from March 25 to April 28, 2020. More than half of the individuals came from the Jianghan District, while others came from different urban and rural areas and different populations in Wuhan. All of them were healthy people with no fever, headache or other symptoms of COVID-19. We aimed to estimate the seroprevalence of immunoglobulin M and G antibodies against SARS-CoV-2 in asymptomatic people in Wuhan, taking into account test performance and adjusting for the age and gender of Wuhan’s population. We also describe the epidemiological characteristics of seroprevalence in its geographic and occupational distribution, as well as chest CT scan and laboratory findings from asymptomatic people. We hope this study will provide a reference for estimating the number of asymptomatic cases of COVID-19 and will improve our understanding of the epidemiology of the outbreak and the development of immunity to COVID-19 in general populations.

## METHODS

### Subjects for serology testing

Before starting the screening for antibodies against SARS-CoV-2, we validated the serological assay with serum samples from 105 patients with COVID-19 confirmed by a SARS-CoV-2 fluorescent PCR kit with nasopharyngeal swabs collected between January 18 and February 22, 2020 (**Supplementary materials-Serology Test Validation**).[24]

Population screening for IgG and IgM against SARS-CoV-2 was initiated on March 25 and was open to all residents of Wuhan who were symptom-free (**Figure S1**). Most of them had returned to work, but a few of them had contact with infected persons who were asymptomatic visiting the designated hospital for COVID-19 diagnosing. The screening and sample collection were carried out in the hospital; hence, the majority of participants resided in the central districts of Wuhan. This was a cross-sectional study to investigate seroprevalence in asymptomatic COVID-19 people in Wuhan for medical examinations. All people lived in Wuhan since the city went into lockdown starting January 22, 2020.

Subject selection criteria: All ages with no fever, headache or other symptoms of COVID-19 in whom specific IgG and IgM antibodies against SARS-CoV-2 were detected. Exclusion criteria: No specific test for SARS-CoV-2 antibodies was completed or clinical data was incomplete. From March 25 to April 28, 2020, a total of 18,712 (89.4%) people met the inclusion criteria with a median age of 40 years (range 4–81 years old), including 11,391 males (60.9%) with a median age of 42 years and 7,321 females (39.1%) with a median age of 37 years.

### Data collection

Clinical data were collected from March 25 to April 28, 2020, including birth date, gender, occupation, residential district, date of test, serum IgG positivity and IgM positivity or negative results for SARS-CoV-2 antibodies, nucleic acid testing, clinical symptoms, chest CT and laboratory tests. Only 1636 of the subjects had chest CTand 12481 subjects had the laboratory tests (**Supplementary materials**). SARS-CoV-2 antibody detection kits (colloidal gold method) were provided by INNOVITA (Tangshan, registration certificate for the medical devices of Peoples Republic of China: 20203400177). All enrolled medical examiners collected 2ml peripheral venous blood. After centrifugation, serum was taken for SARS-CoV-2-specific IgG and IgM antibody detection within 2h. Nasopharyngeal or oropharyngeal swabs used for SARS-CoV-2 nucleic acid testing were collected by trained and qualified medical staff to perform NAT within 2h. The automatic nucleic acid extraction instrument was provided by the Hangzhou Allsheng Instruments Co. Ltd. (Allsheng). The ABI-7500 fluorescence PCR instrument was from Thermo Fisher Scientific Inc. All operations were carried out according to kit instructions.

### Statistical analysis

All analyses were conducted using SPSS software (version 22.0, IBM Corporation, Armonk, NY, USA), the R software package (version 3.6.2; 2019, The R Foundation for Statistical Computing), and GraphPad Prism (Version 8.0, GraphPad Software, LLC, La Jolla, CA, USA). Proportions for categorical variables were compared either by Pearson Chi-square test or by Fisher’s Exact Test. Statistical analysis of continuous variables utilized means with standard deviation (SD) for normally distributed data and medians with interquartile range (IQR) for non-normally distributed data. Means for continuous variables were compared via independent group t-tests when the data were normally distributed; otherwise, the Mann-Whitney test was used. P < 0.05 was considered statistically significant. To correct for the effects of gender and age,[25] the seroprevalence of IgM positivity, IgG positivity, and IgM and/or IgG positivity were standardized according to the gender and age-specific population of Wuhan in 2017 (**Table S1**). To correct for effects from accuracy of the serum antibody test, seroprevalence was corrected according to the sensitivity and specificity of the colloidal gold test in previous studies following the Rogan and Gladen method.[26]

## RESULTS

### Characteristics of individuals studies

Between March 26 and April 28, 2020, we enrolled a total of 18,712 asymptomatic participants, most of them from 154 work units in the central urban area of Wuhan, with a demographic distribution a little different from that of Wuhan (**Figure S2**). The proportion of individuals under 19 and over 65 enrolled in the study was much smaller than the general population. Among those enrolled, 11, 391 (60.9%) were male and 7,321 (39.1%) were female. The median age of the subjects was 40.0 years (IQR, 42–50; range, 4–81), with the majority (n = 17,367, 92.8%) aged 25 to 59 years. The cumulative count of the population tested for antibodies against SARS-CoV-2 and the number of positive cases of IgG and IgM are shown in **Figure 1**.

**Figure 1.**
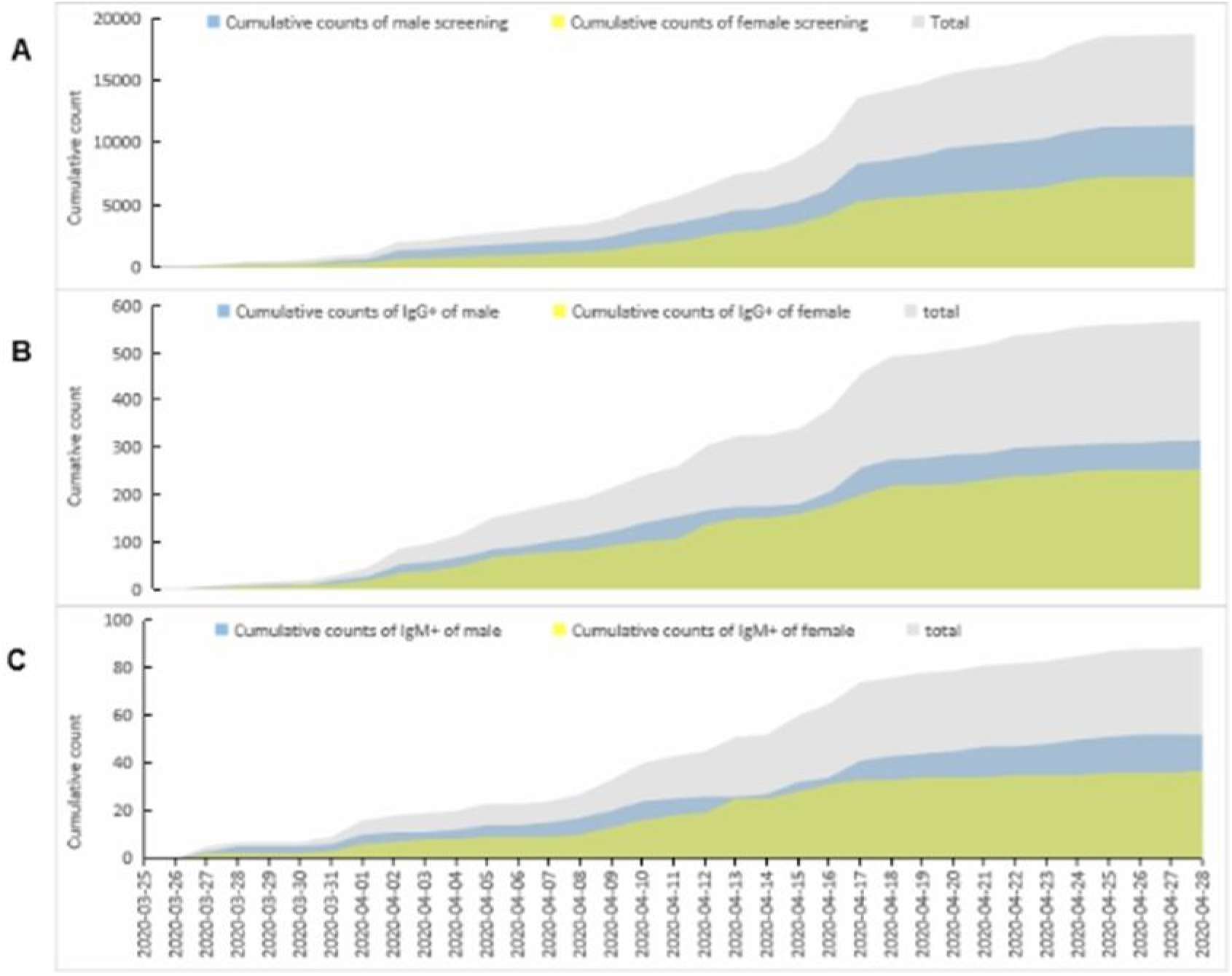
Cumulative count of asymptomatic individuals positive for antibodies against SARS-CoV-2 in a hospital in Wuhan, China, 2020. A: A total of 18,712 asymptomatic individuals tested positive for IgG or IgM antibodies against SARS-CoV-2 from March 25 to April 28, 2020. B: A total of 627 asymptomatic individuals tested positive for serum IgG antibodies, including 351 males and 276 females, from March 25 to April 28, 2020. C: A total of 89 asymptomatic individuals tested positive for serum IgM antibodies, including 52 males and 37 females, from March 25 to April 28, 2020

### Seroprevalence adjusting for age, gender and test

**Table 1** shows the unadjusted and adjusted seroprevalence for IgG and IgM against SARS-CoV-2 in asymptomatic people by gender. The unadjusted seroprevalence of IgG in females was higher than in males and was significantly different (x^2^ =6.525, p = 0.011), resulting in an odds ratio of 1.23 (95% CI: 1.05–1.45). The unadjusted seroprevalence of IgM in females was slightly higher than in males, but was not significantly different (x^2^ =0.225, p = 0.635). When IgG and IgM positivity were analyzed together, the unadjusted seroprevalence of IgG and /or IgM in females was still higher than in males and was significantly different (x^2^ =7.191, p =0.007), producing an odds ratio of 1.09 (95% CI: 1.02–1.17).

**Table 1.**
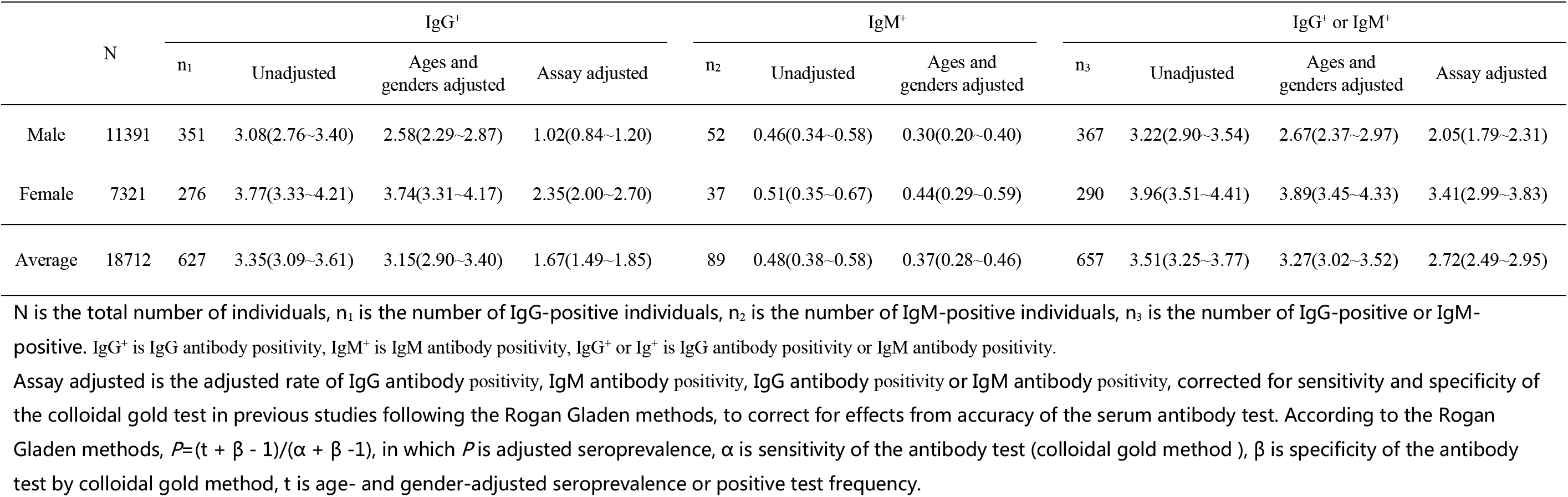
Seroprevalence of IgG, IgM and IgG/IgM in asymptomatic individuals by gender (%, 95% CI)

|  | N | IgG<sup>+</sup> |  |  |  | IgM<sup>+</sup> |  |  | IgG<sup>+</sup> or IgM<sup>+</sup> |  |  |  |
| --- | --- | --- | --- | --- | --- | --- | --- | --- | --- | --- | --- | --- |
|  |  | n<sub>1</sub> | Unadjusted | Ages and genders adjusted | Assay adjusted | n<sub>2</sub> | Unadjusted | Ages and genders adjusted | n<sub>3</sub> | Unadjusted | Ages and genders adjusted | Assay adjusted |
| Male | 11391 | 351 | 3.08(2.76~3.40) | 2.58(2.29~2.87) | 1.02(0.84~1.20) | 52 | 0.46(0.34~0.58) | 0.30(0.20~0.40) | 367 | 3.22(2.90~3.54) | 2.67(2.37~2.97) | 2.05(1.79~2.31) |
| Female | 7321 | 276 | 3.77(3.33~4.21) | 3.74(3.31~4.17) | 2.35(2.00~2.70) | 37 | 0.51(0.35~0.67) | 0.44(0.29~0.59) | 290 | 3.96(3.51~4.41) | 3.89(3.45~4.33) | 3.41(2.99~3.83) |
| Average | 18712 | 627 | 3.35(3.09~3.61) | 3.15(2.90~3.40) | 1.67(1.49~1.85) | 89 | 0.48(0.38~0.58) | 0.37(0.28~0.46) | 657 | 3.51(3.25~3.77) | 3.27(3.02~3.52) | 2.72(2.49~2.95) |

N is the total number of individuals, n<sub>1</sub> is the number of IgG-positive individuals, n<sub>2</sub> is the number of IgM-positive individuals, n<sub>3</sub> is the number of IgG-positive or IgM-positive. IgG<sup>+</sup> is IgG antibody positivity, IgM<sup>+</sup> is IgM antibody positivity, IgG<sup>+</sup> or Ig<sup>+</sup> is IgG antibody positivity or IgM antibody positivity.
Assay adjusted is the adjusted rate of IgG antibody positivity, IgM antibody positivity, IgG antibody positivity or IgM antibody positivity, corrected for sensitivity and specificity of the colloidal gold test in previous studies following the Rogan Gladen methods, to correct for effects from accuracy of the serum antibody test. According to the Rogan Gladen methods, $P = (t + \beta - 1) / (\alpha + \beta - 1)$ , in which $P$ is adjusted seroprevalence, $\alpha$ is sensitivity of the antibody test (colloidal gold method), $\beta$ is specificity of the antibody test by colloidal gold method, $t$ is age- and gender-adjusted seroprevalence or positive test frequency.

After standardization according to the number of people of different ages and gender in the population of Wuhan from the national census of 2017, the age- and gender-adjusted seroprevalence of IgG was higher in females than in males and the difference was significant (x^2^ = 20.433, p < 0.001), with an odds ratio of 1.24 (95% CI: 1.14–1.36). There was no significant difference in IgM between females and males (x2 = 2.436, p = 0.119). The age- and gender-adjusted seroprevalence of IgG and /or IgM was still higher in females than in males, and the difference was significant (x^2^ = 21.906, p < 0.001), with an odds ratio of 1.25 (95% CI: 1.14–1.36).

However, after adjusting for age and gender, and adjusting for test performance (sensitivity and specificity), the overall adjusted serum positive rate for IgG antibodies was 1.67% (95% CI: 1.49–1.85), 1.02% (0.84–1.20) for males and 2.35% (2.00–2.70) for females. After similar adjustments, the overall adjusted serum positive rate for IgG and /or IgM antibodies was 2.72% (95% CI: 2.49–2.95), 2.05% (1.79–2.31) for males and 3.41% (2.99–3.83) for females, and the seroprevalence was significantly higher for females than males (x^2^ = 35.702, p < 0.001) at an odds ratio of 1.36 (95% CI: 1.24–1.48). The census indicated that the Wuhan population aged 4–81 years in 2017 was 7,990,158. Based on this number, asymptomatic COVID-19-positive individuals aged 4-81 years, using IgG and/or IgM seroprevalence tests, were estimated at 217,332 (95% CI: 198,709–235,955) in Wuhan from March 25 to April 28, 2020.

### Distributions in time series and ages

**Table 2** shows the demographic characteristics and seroprevalence of IgG and the odds ratio arranged by time series. In the first five days, we estimated the seroprevalence of IgG at 3.82% (95% CI: 2.18–5.47, n = 523). The estimate increased to 5.14% (4.08–6.19, n=1694) in the second five days, to 8.43% (6.87–9.99, n=1222) in the third five days, and then decreased to3.58% (3.01–4.16, n = 4046) in the fourth five days, to 2.79% (2.40–3.19, n=6695) in the fifth five days, to 2.21% (1.64–2.78, n=2534) in the sixth five days, and finally to 1.45% (0.93–1.98, n = 1998) in the seventh five days period. The positivity for IgM over the same time frame declined slightly and remained at a low level, with the highest rate at 1.33% (0.35–2.32) in the first five days, which dropped to a low of 0.28% (0.07–0.48) in the sixth five days period (**Table S2**).

**Table 2.** Demographic characteristics and seroprevalence of IgG in asymptomatic individuals and odds ratio by time series.

| Five-day period |  | Male, % | Median age in years (IQR) | Crude seroprevalence of IgG | Adjusted seroprevalence of IgG | Crude relative risk (95% CI) | P value | Odds ratio (95% CI) adjusting for age and gender | P value |
| --- | --- | --- | --- | --- | --- | --- | --- | --- | --- |
| 1 | 523 | 55.7 | 38(30~48) | 3.82(2.18~5.47) | 2.98 ( 1.52~4.44 ) | 2.70(1.52~4.81) | 0.001 | 2.62(1.47~4.68) | 0.001 |
| 2 | 1694 | 70.3 | 40(32~51) | 5.14(4.08~6.19) | 4.18 ( 3.23~5.13 ) | 3.68(2.40~5.62) | <0.001 | 3.65(2.38~5.60) | <0.001 |
| 3 | 1222 | 55.3 | 40(31~51) | 8.43(6.87~9.99) | 7.67 ( 6.18~9.16 ) | 6.25(4.11~9.50) | <0.001 | 5.92(3.87~9.05) | <0.001 |
| 4 | 4046 | 61.0 | 40(31~50) | 3.58(3.01~4.16) | 3.18 ( 2.64~3.72 ) | 2.52(1.69~3.77) | <0.001 | 2.42(1.61~3.64) | <0.001 |
| 5 | 6695 | 60.9 | 40(33~49) | 2.79(2.40~3.19) | 2.18 ( 1.83~2.53 ) | 1.95(1.32~2.89) | 0.001 | 1.89(1.27~2.82) | 0.002 |
| 6 | 2534 | 64.0 | 41(31~51) | 2.21(1.64~2.78) | 1.56 ( 1.08~2.04 ) | 1.53(0.98~2.41) | 0.064 | 1.49(0.94~2.36) | 0.086 |
| 7 | 1998 | 55.2 | 32(27~42) | 1.45(0.93~1.98) | 2.23 ( 1.58~2.88 ) | 1(ref) | -- | 1(ref) | -- |

Note: N is number of individuals; IQR is interquartile range.

The distribution of the adjusted seroprevalence of IgG and IgM in the time series, and the distribution of the crude seroprevalence of IgG and IgM in different age groups are in **Figure 2**. The seroprevalence of IgG or IgM in different testing time periods was correlated with the time periods (x^2^=153.88, p < 0.001; x^2^=17.075, p=0 009). Although none of the 26 individuals under the age of 19 years tested positive for IgG, no significant correlation was observed between the seroprevalence of IgG and the different age groups (x^2^ = 11.541, p =0.292). In contrast, the seroprevalence of IgM in different age groups was correlated with age (x^2^ = 18.496, p = 0.035), showing a higher seroprevalence of IgM in the middle-aged group for men and an increased seroprevalence with age for women. No IgM positivity was detected in males or females under 24 years old. The positivity for IgM antibodies in males over 55 years old was significantly lower than for females.

**Figure 2.**
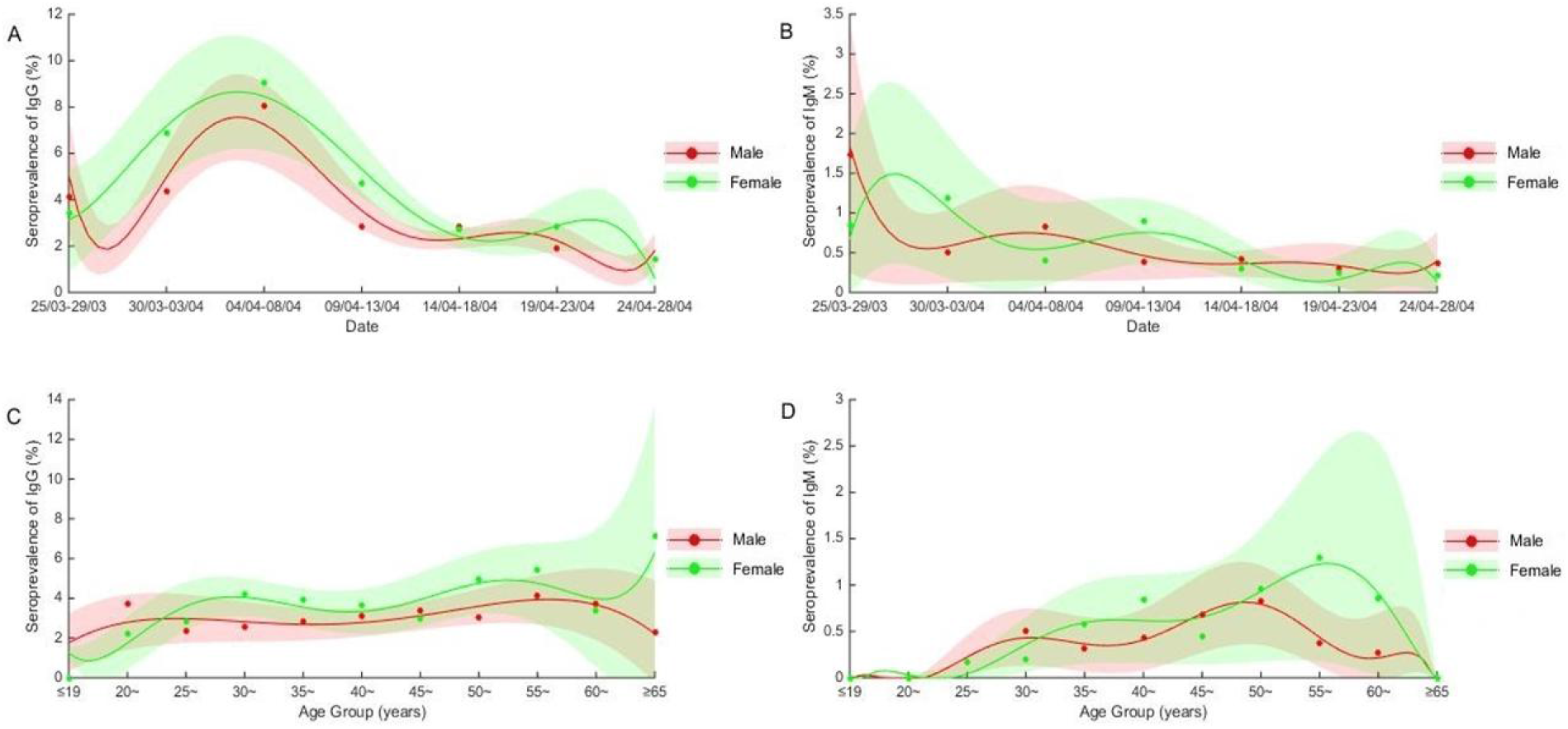
Distributions of seroprevalence of IgG and IgM in asymptomatic individuals in time series and age groups. A: The seroprevalence with 95% CI of IgG adjusted for age in male and female asymptomatic individuals from March 25 to April 28, 2020 in Wuhan, China. B: The seroprevalences with 95% CI of IgM adjusted for age in male and female asymptomatic individuals from March 25 to April 28, 2020 in Wuhan, China. C: The crude seroprevalence with 95% CI of IgG in male and female asymptomatic individuals from ages 4 to 81 in Wuhan, China. D: The crude seroprevalence with 95% CI of IgM in male and female asymptomatic individuals from ages 4 to 81 in Wuhan, China

### Geographic and occupational distribution of seroprevalence

Among the 154 work units, 57.8% (89/154) of work units had at least one positive case of IgG or IgM among employees, accounting for 92.5% (17,315/18,712) of individuals receiving serology testing, including in-service and partly retired employees. Approximately 60% of antibody-positive cases came from the top ten work units having the most number of cases (6.5%, 10/154), including a securities and insurance hall located in Jianghan District with a higher risk for positive cases at 31.0% (9/29), a law firm located in Jiangan District with positive rate of 27.4% (31/113), an urban construction company located in Jianghan District with positive rate of 17.7% (6/34), and rural commercial banks with a positive rate of 7.7% (34/444)). In particular, there was a bank branch with a positive rate of 15.3% (15/98) and two community health service centers with positive rates of 5.06% (9/178), all of them located in Jianghan District. In contrast, there were no positive cases found in serology tests of over 220 individuals from five other community health service centers located far from the central urban area of Wuhan. There were three work units which contributed 30.3% (199/657) of the number of antibody-positive individuals, including a water supply company, a real estate company located in Jianghan District, and telecommunications companies located in Jianghan District.

The crude seroprevalence and odds ratio by geographic areas from different types of workplaces are shown in **Table 3** and **Table 4**. A significant difference was seen in seroprevalence of IgG by urban area (x^2^ = 42.871, p< 0.001), but no significant difference was observed for IgM (x^2^ = 14.729, p = 0.069). The Wuchang and Jianghan districts had higher seroprevalence of IgG than other urban areas. **Figure 3** shows the geographic distribution of the seroprevalence of IgG or IgM antibody against SARS-CoV-2 in asymptomatic people from March 25 to April 28, 2020, in Wuhan, China.

**Table 3.**
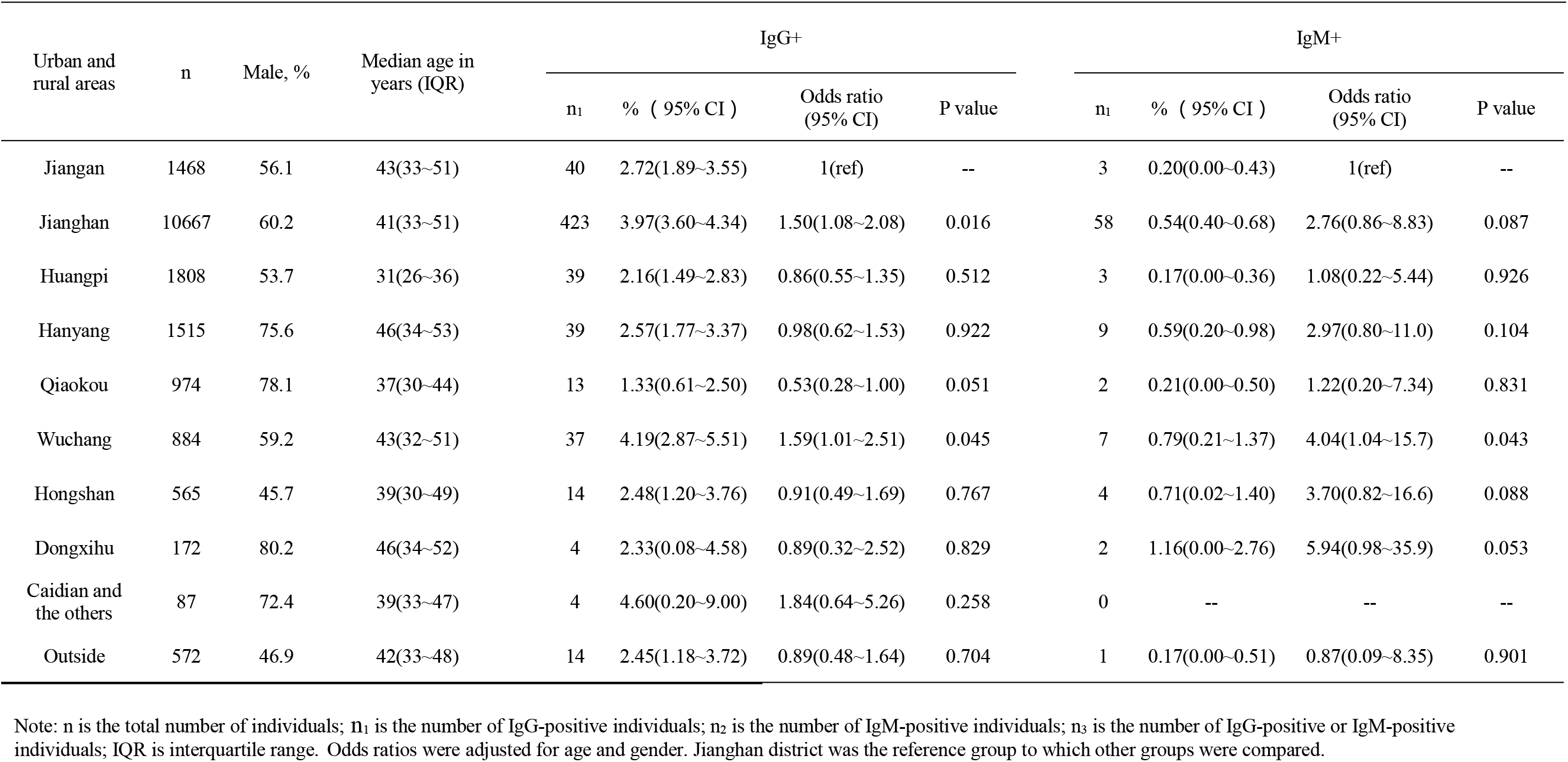
Seroprevalence of IgG and IgM and odds ratio by geographic location (urban and rural areas)

**Table 4.** Odds ratios of the seroprevalence of IgG and IgM by occupation (types of work units)

| Type of work unit | n | Male, % | Median age in years (IQR) | IgG+ |  |  |  | IgM+ |  |  |  |
| --- | --- | --- | --- | --- | --- | --- | --- | --- | --- | --- | --- |
|  |  |  |  | n<sub>1</sub> | % ( 95% CI ) | Odds ratio (95% CI) | P value | n<sub>1</sub> | % ( 95% CI ) | Odds ratio (95% CI) | P value |
| Primary medical care | 628 | 33.9 | 43(34~50) | 15 | 2.39(1.20~3.58) | 1(ref) | -- | 3 | 0.48(0.00~1.02) | 1(ref) | -- |
| Tap water and natural gas | 4961 | 65.6 | 42(35~50) | 96 | 1.94(1.56~2.32) | 0.87(0.50~1.51) | 0.622 | 21 | 0.424(0.24~0.60) | 0.97(0.29~3.29) | 0.961 |
| Real estate | 2481 | 67 | 41(32~52) | 147 | 5.93(5.00~6.86) | 2.78(1.62~4.79) | <0.001 | 17 | 0.69(0.36~1.02) | 1.56(0.45~5.37) | 0.483 |
| Government institutions | 2401 | 66.4 | 44(35~52) | 59 | 2.46(1.84~3.08) | 1.09(0.61~1.94) | 0.762 | 10 | 0.42(0.16~0.68) | 0.92(0.25~3.37) | 0.897 |
| The airport | 1800 | 53.5 | 31(26~36) | 36 | 2.00(1.35~2.65) | 1.01(0.54~1.86) | 0.982 | 3 | 0.17(0.00~0.36) | 0.50(0.10~2.53) | 0.401 |
| Banks, securities and insurance companies | 1786 | 46.7 | 35(30~45) | 89 | 4.98(3.97~5.99) | 2.38(1.36~4.15) | 0.002 | 17 | 0.95(0.50~1.40) | 2.43(0.70~8.37) | 0.161 |
| Highway management and auto repair | 944 | 76.3 | 47(35~54) | 41 | 4.34(3.04~5.64) | 1.99(1.09~3.65) | 0.025 | 8 | 0.85(0.26~1.44) | 1.87(0.49~7.17) | 0.361 |
| Telecommunications companies | 750 | 45.2 | 35(31~40) | 45 | 6.00(4.30~7.70) | 2.96(1.63~5.38) | <0.001 | 1 | 0.13(0.00~0.39) | 0.36(0.04~3.47) | 0.375 |
| Law firms, procuratorates | 706 | 51.4 | 39(31~50) | 40 | 5.67(3.96~7.38) | 2.64(1.44~4.83) | 0.002 | 2 | 0.28(0.00~0.67) | 0.66(0.11~3.98) | 0.652 |
| Rehab | 700 | 72.7 | 45(36~51) | 15 | 2.14(1.07~3.21) | 0.97(0.47~2.00) | 0.925 | 5 | 0.71(0.09~1.33) | 1.62(0.38~6.58) | 0.514 |
| Other service industries | 1501 | 60.4 | 39(31~49) | 33 | 2.20(1.46~2.94) | 1.01(0.54~1.88) | 0.978 | 1 | 0.07(0.00~0.20) | 0.16(0.02~1.53) | 0.112 |
| Personal | 54 | 61.1 | 42(32~49) | 11 | 20.4(9.63~31.1) | 11.5(4.95~26.6) | <0.001 | 1 | 1.85(0.00~5.44) | 4.40(0.45~43.3) | 0.204 |

Note: n is the total number of individuals; n<sub>1</sub> is the number of IgG-positive individuals; n<sub>2</sub> is the number of IgM-positive individuals; n<sub>3</sub> is the number of IgG-positive or IgM-positive individuals; IQR is interquartile range. Real estate includes real estate agencies, residential property management firms and urban construction corporations. Odds ratios were adjusted for age and gender. Primary medical care was the reference groups to which other groups were compared.

**Figure 3.**
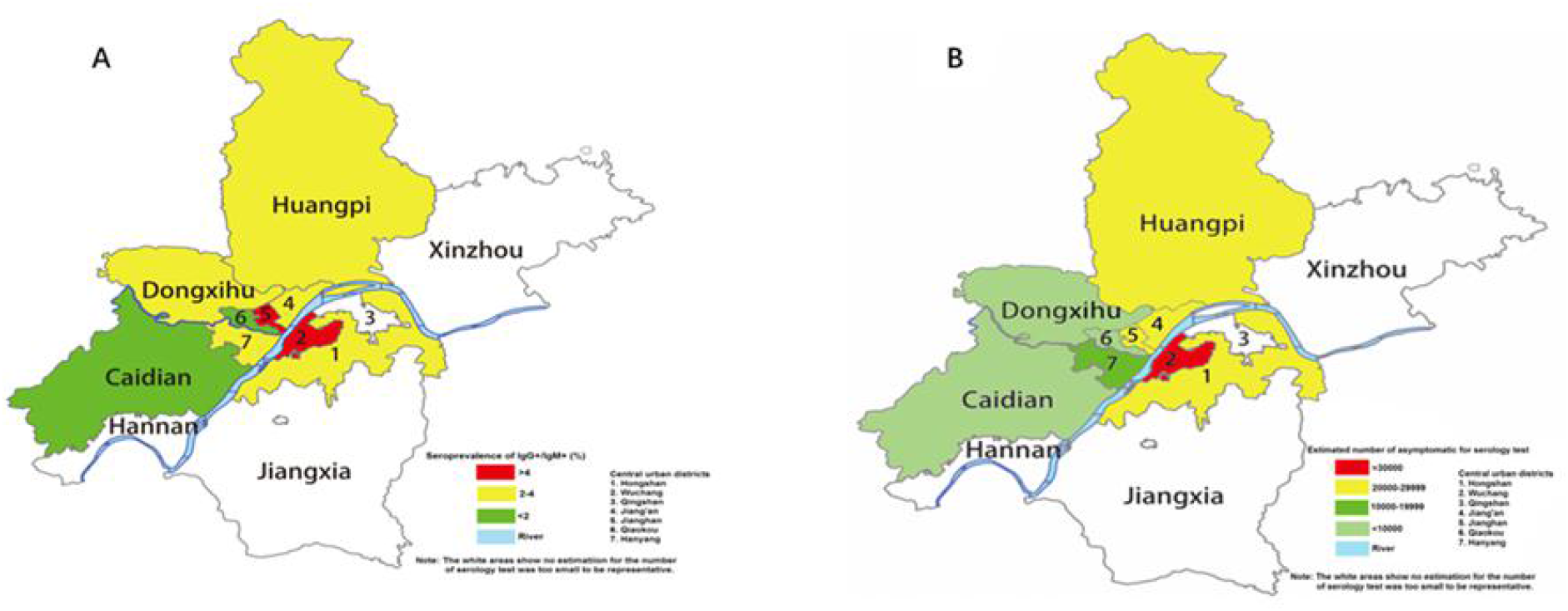
Geographic distribution of asymptomatic people in Wuhan. A: The geographic distribution of seroprevalences of IgG or IgM antibodies against SARS-CoV-2 in asymptomatic people from March 25 to April 28, 2020, in Wuhan, China. B: The geographic distribution of asymptomatic people estimated according to the seroprevalences and residential populations in the districts of Wuhan in 2017.

**Figure 4.**
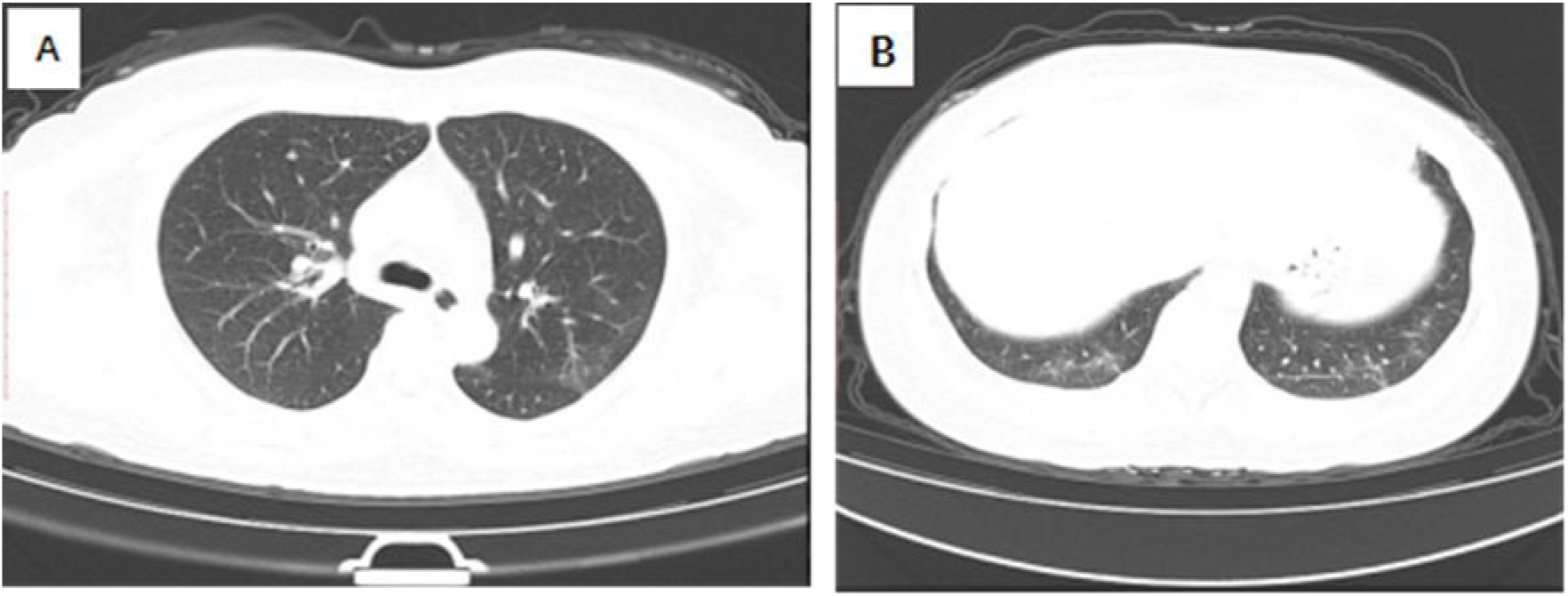
CT imaging of one asymptomatic individual positive for IgG. A shows a platelet of GGO in the posterior segment of the upper lobe of the left lung. B shows GGOs with small mixed consolidation in the posterior basal segment of the lower lobe of both lungs, and the pleural offline was seen in the posterior basal segment of the lower lobe of the left lung.

A significant difference was seen in the seroprevalence of IgG and IgM among people from different work units (x^2^ = 202.43, p < 0.001 for IgG; x^2^ = 28.915, p = 0.001 for IgM). Among the tested individuals, the seroprevalence of IgG was highest for those who voluntarily came for serology test by themselves, followed by those encouraged by their work units, including real estate property companies, urban construction companies, telecommunications companies, banks, securities and insurance companies, law firms, and procuratorates.

### CT Imaging and laboratory findings

The relationship between abnormal chest CT imaging and the seroprevalence of IgG and IgM antibodies for different age groups is shown (**Figure S3 and Figure S4**). Of the participants, 1636 underwent chest CT, and 342 had abnormal images. CT imaging abnormality was 30.7% (94/306) for people who were IgG antibody-positive compared to 19.7% (262/1330) for those who were IgG negative. The difference was significant (x^2^ = 17.743, p < 0.001), as shown by the odds ratio of 1.13 (95% CI: 1.06–1.21). Among the IgG-positive patients, typical abnormal chest CT signs were GGOs, GGOs with small mixed consolidation, and prominent peripheral subpleural distribution. Significant differences were seen between IgG and/or IgM-positive test groups in leucocytes, neutrophilic granulocytes and monocyte percentage (**Table S4**).

## DISCUSSION

Detection of asymptomatic or subclinical novel human coronavirus SARS-CoV-2 infections is critical for understanding the overall prevalence and infection potential of COVID-19.[9] There were some SARS-CoV-2 seroprevalence studies of target people earlier this year from blood donors or outpatient settings,[15-16] and from the general population of China,[10-12] theUSA,[13] Switzerland,[14] and Spain.[15] The Switzerland study showed seroprevalences from 4.8% to 10.9% in different weeks in Geneva. The Spanish study showed a nationwide seroprevalence of 5.0% (4.7–5.4%), with urban areas around Madrid exceeding 10%. All of these studies demonstrate that most of the population appears to have remained unexposed to SARS-CoV-2. Our study aimed to estimate the seroprevalence from SARS-CoV-2 infections in Wuhan, the epicenter of the COVID-19 pandemic in China. Despite being the earliest area to be affected by COVID-19, the seroprevalence among the general population was only 3.35%. Our study indicates that most of the Wuhan population remains susceptible to COVID-19. There were 50,333 reported confirmed patients up to April 28 in Wuhan, and we estimate that for every reported confirmed case, there were at least 4.1 (204,548/50,333) asymptomatic infections in the general population. This is smaller than the estimate of the Switzerland study, which estimated 11.6 infections in the community for every reported confirmed case.

Our study showed there were differences in IgG seroprevalence among urban and rural areas, which were much higher in Wuchang District and Jianghan District than in other areas. Based on the cumulative number of reported cases of COVID-19 from March 25 to April 28, and the number of residents in each district in Wuhan, we estimated an average incidence rate per 100,000 people of 2.98% (95% CI: 2.97–2.99) during this period. Incidence rates were higher in the Jianghan and Wuchang districts, at 8.09% (95% CI: 8.02–8.15) and 7.25% (95% CI: 7.20–7.29) per 100,000 people, respectively, which show that seropositive rates in different geographic areas were consistent with the spread of the SARS-CoV-2 coronavirus in Wuhan (**Table S3 and Figure S5**).

In our study, the highest seroprevalence of IgG was in those people who voluntarily came for serology testing by themselves, most of whom had close contacts with COVID-19 patients and therefore had a higher risk of infection. Our study found significant aggregation of asymptomatic infections in individuals from certain occupations. The groups in some industries with high IgG seroprevalence were easy to identify due to the nature of their jobs, such as insurance and securities professionals, professionals in the law industry, maintenance personnel of urban construction company, and staff of rural commercial banks located in the higher risk districts. It is known that some staff undertook substantial voluntary work in response to government calls during the COVID-19 pandemic, to assist in, for instance, administering in the local residential communities, transporting COVID-19 patients, and screening the general population for epidemic incidence. Thus, they faced a greater chance of contacting infected people during their work, and were more likely to be infected, even with personal protection.

Some studies found an average level of IgG antibodies higher in females than in males.[27] In our study, the IgG seroprevalence was also higher in females than in males, indicating that females were more likely to have asymptomatic infections.

To our knowledge, the most common CT features in patients affected by COVID-19 include GGO and consolidation involving the bilateral lungs in a peripheral distribution.[28] In our study, CT imaging demonstrated similar differences between IgG antibody-positive and negative individuals, which is a reminder that some damage may have occurred in some asymptomatic individuals positive for IgG antibodies.

This study had a few limitations. First, this study had selection bias since the analyzed medical records were based on examinees directed by their work units. Most of the examinees came from government-owned institutions and agencies instead of private businesses. Therefore, the sample was incompletely randomized and insufficiently representative, compromising the assessment accuracy of the prevalence of asymptomatic infections in Wuhan. Second, as the examinees were only from the back-to-work population, people under the age of 19 and over age 65 were too few to be fully covered in analyses.

This study has the important feature of having been designed as repeated five-day serosurveys, which allowed for the monitoring of seroprevalence progression just after the end of the first epidemic. Further, our study applied scientific statistical methods accounting for the demographic structure of the general population and imperfect diagnostic tests to estimate seroprevalence in the overall population, while capturing uncertainty in the estimates. Our study will provide useful information for the investigation of herd immunity and help design targeted strategies for prevention. Additionally, the study of IgG and IgM against SARS-CoV-2 among asymptomatic infections can provide scientific basis for vaccine development.

## CONCLUSIONS

The seroprevalence of IgG and IgM in Wuhan indicates that reported confirmed patients only represent a small proportion of the total number of infections, and most of the Wuhan population remains susceptible to COVID-19. There were differences in IgG seroprevalence among geographic areas, which were consistent with the spread of the SARS-CoV-2 coronavirus in Wuhan. Our study found significant aggregation of asymptomatic infections in individual from certain occupations. The CT imaging and laboratory tests suggest that some slight damage may have occurred in some asymptomatic individuals positive for IgG antibodies.

## Data Availability

All data referred to in the manuscript is availability.

http://wjw.wuhan.gov.cn/ztzl_28/fk/tzgg/202004/t20200430_1202887.shtml

http://wjw.wuhan.gov.cn/ztzl_28/fk/tzgg/202004/t20200430_1198159.shtml

http://tjj.wuhan.gov.cn/tjfw/tjfx/202001/t20200115_840940.shtml

## Acknowledgements

We acknowledge the work and contribution of all the health care providers from the Hubei Provincial Hospital of Integrated Chinese & Western Medicine.

## Ethical approval and consent to participate

This study was approved by the Ethics Committee of the Hubei Provincial Hospital of Integrated Chinese & Western Medicine (No. 2020011). Waiver of informed consent for collection of epidemiological data from individuals tested for COVID-19 was granted by the National Health Commission of China as part of the infectious disease outbreak investigation. All identifiable personal information was removed for privacy protection when data were extracted.

## Author Contributors

RJL, YHY, HQW and TL were responsible for study design. RJL, YHY, HQW and JYH were responsible for the literature search. RJ, YHY, JXZ, SX, RRS, WCZ, HQW and MFC were responsible for data collection. HQW was responsible for data analysis. HQW and MFC were responsible for figures. HQW, HLJ and TL were responsible for data interpretation. HQW, JYH, HLJ, RJL, and YHY were responsible for writing the first draft of the manuscript. All authors contributed to the final draft.

## Patient and Public Involvement

Patients or the public WERE NOT involved in the design, or conduct, or reporting, or dissemination plans of our research.

